# Construction of a risk prediction model of postoperative delirium in older patients undergoing spinal surgery: Protocol of a single-center observational cohort study

**DOI:** 10.1101/2023.04.10.23288343

**Authors:** Qian Liu, Meng Jiao, Ke Huang, Xuexin Feng, Tianlong Wang

## Abstract

**Background:** Delirium is an acute mental disorder and a common postoperative complication. Age is an independent marker of the development of postoperative delirium. In older patients, delirium leads to numerous detrimental effects. We will mainly explore the incidence and potential risk variables of postoperative delirium in older patients undergoing spinal surgery, focusing on some preoperative and intraoperative indicators.

**Study design and methods:** This single-center prospective, observational, cohort study will investigate the incidence of delirium in patients aged ≥65 years undergoing spinal surgery and construct a postoperative delirium risk prediction model. We will use potential multiple risk factors reported in recent studies. Follow-up starts on the first day after the operation, and delirium assessment is conducted until 7 days after the operation. The least absolute shrinkage and selection operator regression will be used to filter variables, and logistic regression will be utilized to build a prediction model using the selected variables. The area under the receiver operating characteristic curve will be used to evaluate the accuracy of the prediction model. The clinical net benefit of the model will be evaluated using decision curve analysis.

**Discussion:** This study will construct a clinically effective model to predict the occurrence of postoperative delirium in older patients undergoing spinal surgery.

## 1. Background

Postoperative delirium is a complex neuropsychiatric syndrome with a high prevalence in older patients and is associated with adverse outcomes, including severe morbidity and mortality. Postoperative delirium is also known as acute brain dysfunction, and the disease course is characterized by a disruption of the patient’s state of consciousness, perception, orientation, memory, cognition, and psychomotor behavior within a week after surgical surgery^[1]^. As one of the common postoperative complications, postoperative delirium occurs in 10%–55% of patients. Delirium is associated with poor prognosis, leading to an increased risk of death, prolonged recovery time, longer hospitalization, increased hospitalization expenses, and increased development of long-term cognitive dysfunction^[2]^. Therefore, reducing the incidence of postoperative delirium by providing appropriate interventions is urgent.

With the progress in medical technology and the improvement of living standards, an increasing number of older patients receive surgical treatment, and related postoperative complications also increased. In China, people aged >60 years account for nearly one-fifth of the total population and have reached 249 million in 2018. People aged >60 years will reach close to 450 million and >30% of the global population by 2050^[3, 4]^.

Spinal surgery in older patients represents a major public health concern, i.e., difficulty in achieving good postoperative outcomes including delirium, and subsequently adds a heavy socioeconomic burden on society. Comprehensive application of various preventive means cannot only reduce the risk of delirium but also reduce its duration and severity and improve the prognosis of older patients^[5]^. Thus, early recognition of the risk factors of delirium is crucial.

This prospective observational single-center cohort study aims to construct a delirium prediction based on multiple scores and determine the incidence of delirium in older patients undergoing spinal surgery.

## 2. Study Design, Setting, and Population

### 2.1 Study design

This is a prospective, single-center, observational, cohort study.

### 2.2 Setting

All investigations will be conducted in conformity with the ethical principles of research. The study was overseen and approved by an independent institutional internal review board and ethics committee (Identifier IRB-2022-202) on 12/02/2022. The trial was also registered in the China Clinical Trial Center (Identifier ChiCTR-2300068660) on 02/28/2023. The participating institution complies with the Declaration of Helsinki, and this is an observational study without interventions. All patient data will be collected anonymously and recorded to ensure patient confidentiality.

For perioperative treatment and surgical procedures, our hospital has specialized geriatric spinal surgery wards. The assessing team is composed of one internal medicine doctor who is responsible for patients’ perioperative management, two orthopedists, an attending anesthesiologist, and nurses.

### 2.3 Population

Inclusion criteria

➢ Age ≥65 years, no sex and nationality limitations
➢ American Society of Anesthesiologists (ASA) grades I–III
➢ Elective spinal surgery as the surgery type
➢ Acceptance of general anesthesia
➢ Consent to participate in this study and sign the informed consent form
➢ Complete case data and scheduled operation

Exclusion criteria

➢ Emergency surgery
➢ Refusal to participate in the study
➢ Lack of cooperation or communication abilities, such as the following conditions:
  - Allolalia
  - Dysaudia
  - Coma
➢ Patients are unable to read Chinese
➢ Epilepsy
➢ Parkinson’s disease
➢ Dopamine drugs (levodopa and dopamine agonists)
➢ Severe condition
➢ ASA grade ≥IV.

Discharge criteria

➢ Discharge without surgical treatment
➢ Withdrawal from the study at any time
➢ Secondary surgery during the follow-up period
➢ Loss of the data
➢ Lost to visit
➢ Participants withdrew actively
➢ Violations of test procedures

## 3. Anesthesia and Data Collection

### 3.1 General baseline data

Factors that maybe potential variables for postoperative delirium include age, sex, height, weight, body mass index, general health status based on the ASA classification, preoperative pain score on the visual analog scale (VAS), educational level, residence (rural or urban), history of food and drug allergy, surgery and anesthesia for the 2 weeks before enrollment, smoking (defined as continuous or cumulative smoking for ≥6 months in the lifetime and those who have not stopped smoking for two consecutive years), alcoholism [definite daily intake of pure alcohol of >25 g (equivalent to 750 mL of beer, 250 mL of wine, or 75 mL of 38% liquor, or 50 mL of 5% liquor), daily intake of pure alcohol of >15 g in women (equivalent to 450 mL of beer, 150 mL of wine, 50 mL of 38% liquor, or 30 mL of 50% liquor)], frailty, anxiety/depression, cognitive impairment, cognitive reserve, preoperative application of psychotropic drugs (benzodiazepines, anticholinic, or antihistamines), history of multiple medications (>5 drugs), history of stroke, malnutrition, visual/hearing impairment, sleep disorders, dehydration [blood urea nitrogen (mg/dL)/creatinine (mg/dL) 18 (except for fever, high-protein diet, taking steroids, gastrointestinal bleeding, and cardiac insufficiency)], anemia, hypoproteinemia, preoperative hypoxemia, electrolyte abnormalities, and spin disease type. All factors will be recorded after enrollment.

### 3.2 Data collection on the preoperative laboratory

The following data will be collected: routine blood examination (white blood cells, neutrophils, red blood cells, hemoglobin, and platelet count), biochemical examination (electrolytes, alanine transaminase, glutamate aminotransferase, total bilirubin, total protein, albumin, lactic acid, dehydrogenase, blood urea nitrogen, and serum creatinine), and routine urine examination before entering the operating room.

### 3.3 Assessment of preoperative physical and psychological state

For an accurate and comprehensive evaluation of coexisting disease index, weakness, mental, psychological state, nutritional status, sleep status, ability to perform daily living activities, and social support will be collected. A series of evaluation indices are will be used to evaluate the physiological status preoperatively, including the age-adjusted Charlson comorbidity index(aCCI), FRAIL scale, simplified mini-mental state examination(MMSE scale), Hospital Anxiety and Depression Scale (HAD), Social Support Rating Scale (SSRS)^[6]^, modified PAP index evaluation form (modified Barthel index), and Pittsburgh sleep quality index. The electrocardiogram test results will be collected 3 days before enrollment, and imaging results will be the data available for the week before enrollment.

### 3.4 Assessment of anesthesia and intraoperative data

The operation time, anesthesia duration, bleeding amount, blood transfusion amount, intraoperative hypotension, intraoperative hypertension, intraoperative cardiac arrhythmia, and recovery from anesthesia will be collected from the electrical anesthesia systems.

### 3.5 Postoperative data and follow-up

The assumption of sedation medicine, assumption of analgesia, anticholine, postoperative destination, postoperative pain score (VAS), other serious complications requiring treatment (including myocardial infarction, heart failure, deep vein thrombosis, pulmonary embolism, infection, liver and kidney dysfunction, and stroke), hospital stay, and postoperative hypoxia will be recorded after the operation.

The follow-up will start from the first postoperative day, once a day, until 7 days after surgery and 1 week after discharge. The occurrence will be evaluated using the 3D confusion assessment method (CAM). If the patient enters the intensive care unit (ICU) after surgery, the use of the CAM-ICU starts on the first postoperative day and continues until 7 days.

### 3.6 Quality control

Before the start of the formal trial, the researcher shall formulate the clinical research plan and submit it to the ethics committee for approval; conduct unified training for all participants, including training on the use of evaluation scales, unified recording methods, and judgment standards. During the study, the sponsor should regularly conduct on-site supervision visits to the study implementers to ensure that all contents of the study plan are strictly followed, and the information filled in is correct. All observation results and findings should be verified to ensure data reliability and ensure that all conclusions in the clinical validation come from the original data. Researchers should objectively and truthfully record and retain all data and protocol implementation and modification during the research process. At the stage of patient recruitment, the consistency of inclusion/exclusion criteria should be guaranteed as much as possible.

## 4. Outcome Measures

The primary outcome is the incidence and related risk factors of delirium following spinal surgery to establish a predictive model using potential variables.

## 5. Sample Size Calculation

The sample size is calculated according to the two-category outcome formula of the clinical predictive model recommended by Riley^[7]^. PASS V.11 software (NCSS, Kaysville, Utah, USA) is used. Combined with previous studies that reported a 20% incidence of delirium after spinal surgery and our clinical experience, based on 90% power to detect a significant difference (α = 0.05, two-sided) the sample size is approximately 876. Considering the dropout rate of 10%, finally, 964 cases will be selected for this study.

## 6. Statistical Management and Data Monitoring

A monitoring committee will be established to control data accuracy. A member of the study team will be appointed to monitor the whole research regularly. Patients’ demographic, clinical, and perioperative data must be checked by two researchers. A researcher will review the medical records of all included patients and collect perioperative data. Meanwhile, 5% of all patients will be randomly selected by another researcher, who will independently confirm the accuracy of the collected data.

If any discrepancy occurs, it will be decided by the third researcher.

Statistical analysis will be performed using the R software package. Data with normal distribution will be presented as mean ± standard deviation, and the t-test will be used for further analysis. For data without normal distribution, they will be presented as median and interquartile range and will be analyzed using nonparametric tests. Data classification will be based on the frequency, relative number (rate or composition ratio) description using the chi-square test or Fisher test. *P* < 0.05 was considered statistically significant.

The least absolute shrinkage and selection operator will be used to screen related influencing factors, and these selected variables will be further analyzed by logistic regression. The area under the receiver operating characteristic curve will be used to describe the calibration degree with the calibration curve. To evaluate the clinical benefit of the model, the decision curve analysis will be employed. The model will be internally verified by the Bootstrap method and externally verified by the period verification method.

## 7. Discussion

Delirium is a common complication that afflicts 20%–80% of older patients undergoing surgery^[8]^. Although delirium is a mental psychological symptom, multiple preoperative somatic factors can promote the occurrence of delirium postoperatively^[9]^, such as advanced age, malnutrition, dehydration, reduced physiological function and cognitive reserve, stroke history, comorbidities^[2, 9]^. With the development of spinal surgery technology and the improvement of the anesthesia level, many older patients with severe spinal diseases have undergone surgery and achieved good postoperative results. However, some older patients have abnormal mental status after surgery, such as anxiety, complacency, agitation, and other states, which increase the difficulty and risk for recovery and treatment during the perioperative period. Owing to the decline in physiological function and different severity of comorbidities, older patients are more susceptible to risk factors of delirium. A meta-analysis of postoperative risk factors reported that the incidence of delirium varies with surgeries, such as 14.7% after head and neck surgery, 19.1% for vascular surgery, 22.2% for gastrointestinal surgery, 24% for hip fracture surgery, 54.9% for cardiac surgery, and 41% for spinal surgery^[10-15]^. Focusing on identifying high-risk patients preoperatively, approximately 40% of the cases are preventable^[16]^.

In the past few decades, researchers have constructed numerous postoperative risk prediction models. Dai^[17]^ reported data of patients with orthopedic and urology problems (mean age, 72.7 years). Risk factors were divided into baseline factors (advanced age and cognitive impairment) and predisposing factors (use of psychotropic drugs). The model included five risk groups: very low risk (no risk factor), lower risk (one risk factor), low risk (two baseline factors), medium risk (one induction factor and one baseline factor), and high risk (three risk factors), with predicted rates of 0.5%, 6.5%, 5.6%, 20.6%, and 28.1%, respectively. Min Young^[18]^ studied patients with general surgical procedures, and nine risk factors, namely, age, low mobility, alcohol abuse, hearing impairment, previous medical history, emergency surgery, open surgery, ICU admission, and elevated C-reactive protein(CPR), were included for the Delphi (delirium prediction based on hospital information) score. With a maximum score of 9 points, those who scored 7 points will be high-risk patients after surgery. The sensitivity and specificity of the model are approximately 80.8% and 92.5%, respectively. The AWOL scoring system^[19]^ includes five risk factors, namely, age ≥ 80 years, inability to spell the word “world” in reverse order, disorientation, serious complications, and high-risk surgery. The predicted risk was automatically calculated by the system, and ≥5% is defined as high-risk patients. The model has sensitivity and specificity of approximately 75% and 59%, respectively, with moderate accuracy. However, it does not apply to nonnative English-speaking countries.

Degenerative lesions in spinal surgery are commonly prevalent in older patients. In recent years, an increasing number of older (75 years old) patients or even super old (85 years old) patients have received spinal surgical treatment. Preoperatively, patients have different degrees of functional restriction, such as numbness, distension, pain, weakness, difficulty in performing complete fine movements, lower limb weakness, and intermittent claudication. Therefore, some scholars have constructed some prediction models for this kind of surgery. A single-center study was conducted in patients with a mean age of ≥75 years and showed that the examination of frailty and cognitive function with the FRAIL scale and animal fluency test, respectively, could rapidly screen postoperative high-risk populations before surgery^[20]^. Among them, frailty (FRAIL score of 3 points) was a strong predictor of postoperative delirium (odds ratio [OR] 6.6). Low scores on the animal fluency test (OR 1.08) and more invasive surgical procedures (OR 2.69) were also associated with postoperative delirium. Another recent study suggested that preoperative cognitive impairment (OR 2.45), depression (OR 4.54), and complex surgery (OR 5.88) were associated with higher incidence of postoperative delirium. The prediction accuracy of FRAIL was 72%, which was significantly higher than the surgery-specific risk (AWOL-S) model (prediction accuracy of 45%)^[20]^. The above two models lack relevant validation and are not easy to use within the ward. Owing to these limitations, a convenient and effective evaluation system for older patients undergoing spinal surgery is necessary. This study intends to construct and verify a prediction model for the risk of postoperative delirium in older patients undergoing spinal surgery in China and determine the incidence of postoperative delirium to provide clinical guidance.

In summary, this prospective, single-center, observational cohort study aims to construct a risk prediction model of postoperative delirium in older patients undergoing spinal surgery. This trial will reveal the key risk factors of postoperative delirium and will provide important clinical guidance for early geriatric delirium detection, assessment, and management.

**Figure 1.**
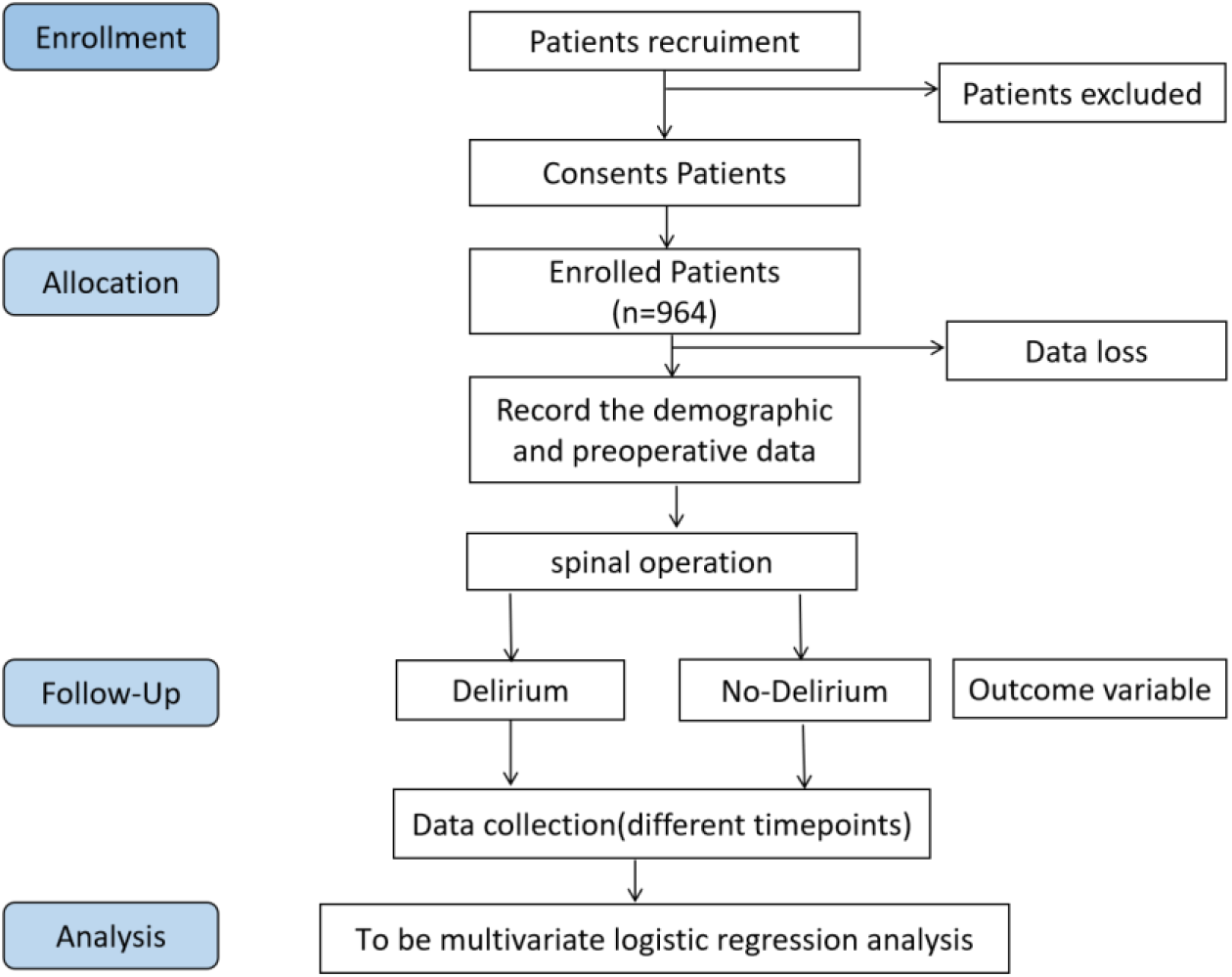
Flowchart of the study procedure.

**Table 1.**
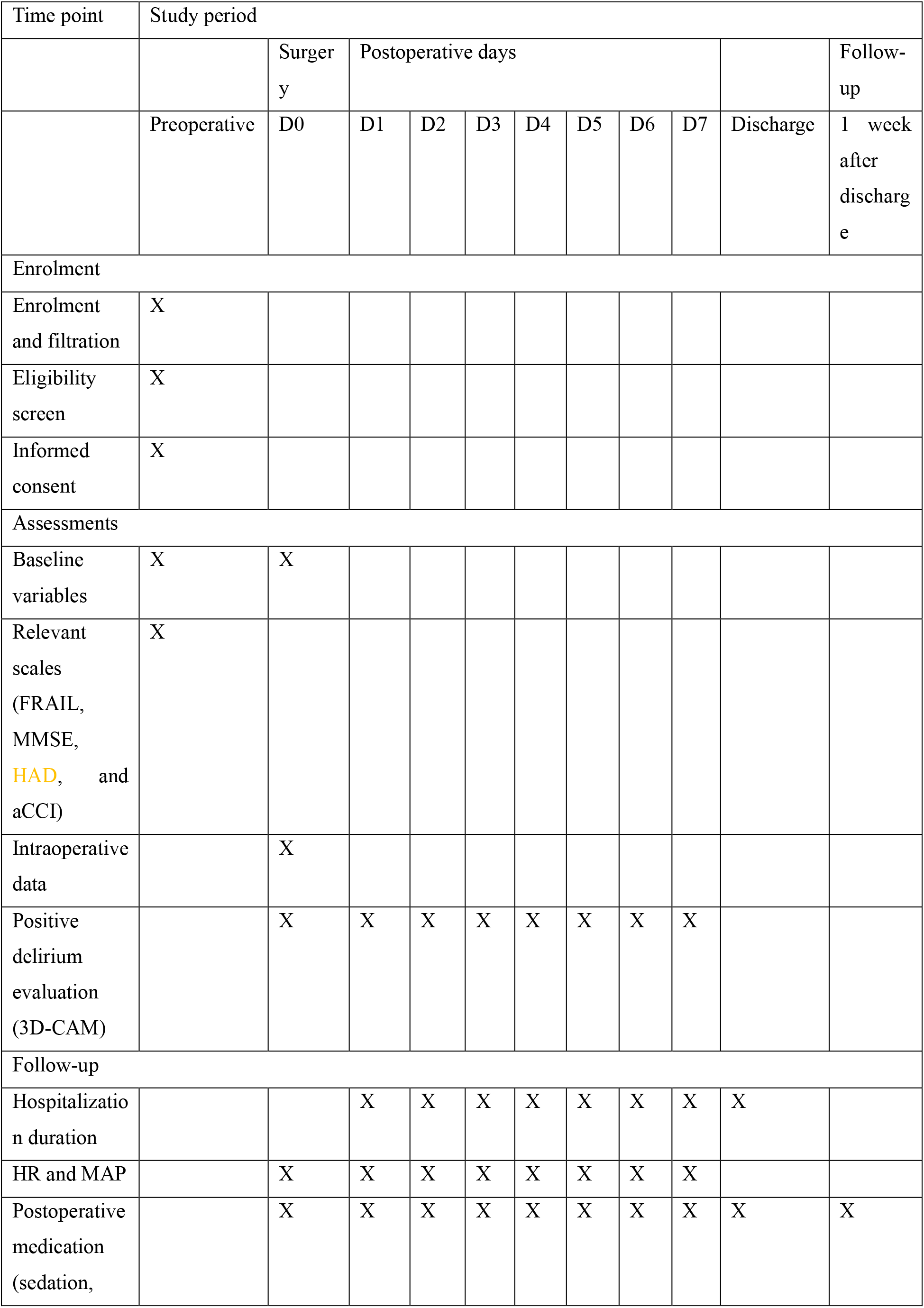
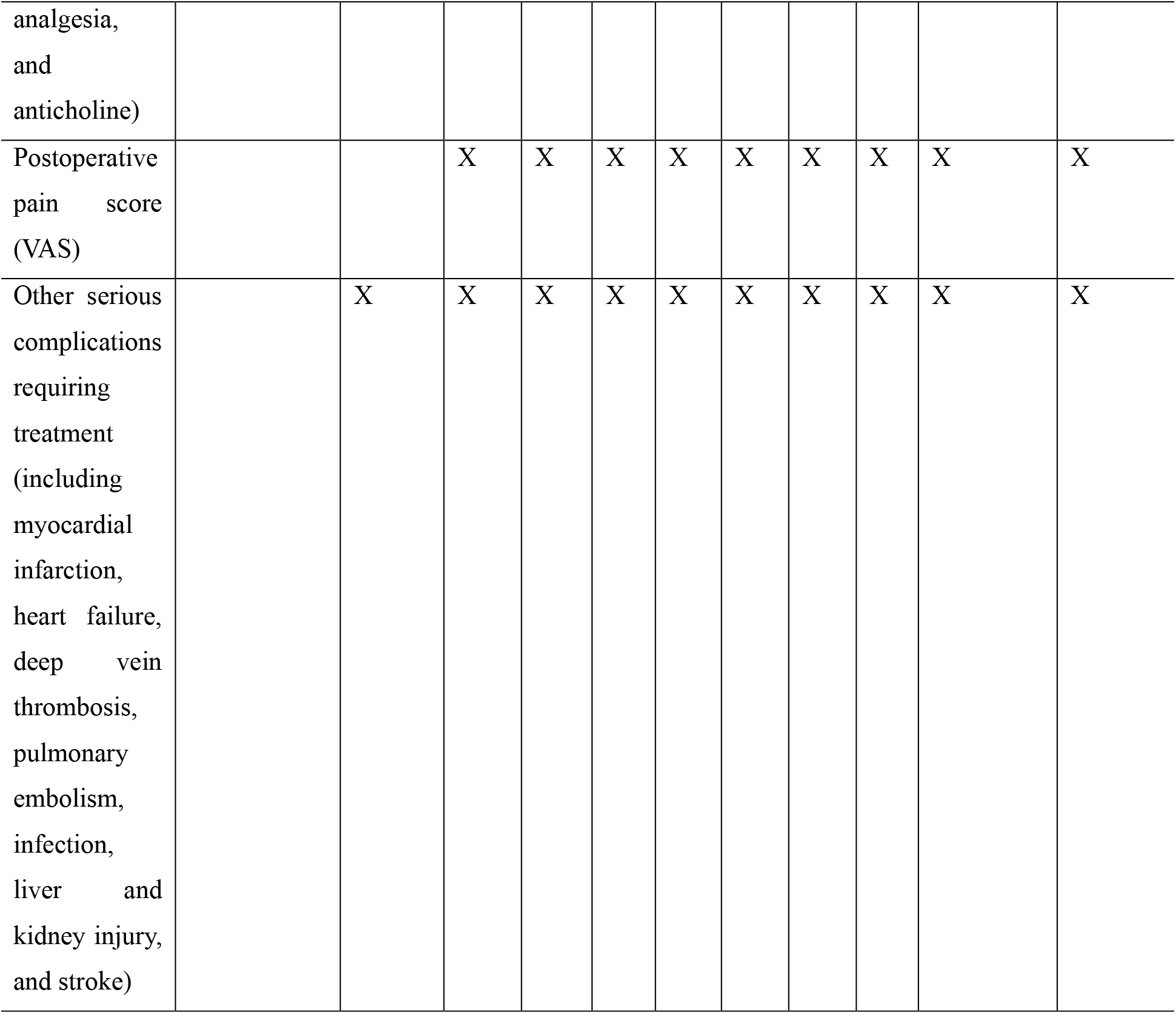
Trial schedule

## Data Availability

All data produced in the present study are available upon reasonable request to the authors

